# Pregnant women’s knowledge of obstetrical danger signs: A cross-sectional survey in Kigali, Rwanda

**DOI:** 10.1101/2022.05.03.22274645

**Authors:** Emmanuel Uwiringiyimana, Emery Manirambona, Samuel Byiringiro, Albert Nsanzimana, Neophyte Uhawenayo, Pacifique Ufitinema, Janviere bayizere, Patricia J. Moreland, Pamela Meharry, Diomede Ntasumbumuyange

## Abstract

**BACKGROUND:** Maternal mortality remains critically high worldwide, particularly in sub-Saharan Africa. The leading causes of maternal death in Rwanda include postpartum hemorrhage and obstructed labor. Maternal recognition of obstetrical danger signs is critical for timely access to emergency care, in order to reduce maternal mortality.

**OBJECTIVE:** To assess the knowledge of obstetrical danger signs among pregnant women attending antenatal care in Kigali, Rwanda.

**METHODS:** A cross-sectional study was conducted between September and December 2018 at five health centers and one district hospital in Kigali, Rwanda. Pregnant women attending antenatal care (ANC) services completed a structured questionnaire. Data were analyzed using descriptive statistics.

**RESULTS:** A total of 382 pregnant women were included in the study. The majority of women (67.8%) were aged 23-35 years, and 44.5% had completed secondary education. Almost half (43.2%) reported traveling more than 30 minutes to reach the health facility; only 23.3% were within 15 minutes of the health facility. Over half (57%) reported attending three or more ANC visits during pregnancy. The majority (85.6%) knew at least one obstetrical danger sign, with nearly half (46.1%) obtaining knowledge of danger signs from midwives and nurses.

**CONCLUSION:** Knowledgeability was significantly associated with the parity and number of ANC visits, though CHW was also a good source of information for pregnant women. We encourage a systematically designed curriculum to teach mothers during their follow-up visits for ANC.

## INTRODUCTION

The complications from pregnancy and childbirth are the leading causes of maternal deaths worldwide (1–3). The burden of maternal mortality is greatest in low- and middle-income countries (LMICs), with 67% occurring in Sub-Saharan Africa (SSA), and 27% in Southern Asia (4). Half of all global maternal deaths result from post-partum hemorrhage (27%), hypertensive disorders (14%), and sepsis (11%)(1). Evidence shows that most maternal complications during pregnancy are preventable via timely recognition of danger signs and effective interventions at health facilities (4). The danger signs during pregnancy and after delivery are the warnings of imminent or ongoing life-threatening events requiring quick intervention by skilled healthcare providers(5). In studies conducted in Ethiopia, Tanzania, and Uganda less than 60% of respondents had knowledge of at least 3 danger signs. Evidently, maternal knowledge of obstetrical danger signs (ODS) is very low in SSA, which prevents early care seeking by mothers, and contribute to a higher maternal mortality (6–9). To achieve the Sustainable Development Goal 3 (SDG3) target of reducing maternal deaths to less than 70 per 100,000 births by 2030, strategies will have to include the improvement of maternal knowledge of ODS (7,10).

Several factors predict women’s knowledge of ODS and could provide guidance on where to target interventions. According to a study conducted in Ethiopia, the attendance of Antenatal Care (ANC) was associated with 1.26 times higher knowledge, and women who delivered at the health facility were 3.57 times more likely to be knowledge of ODS (11). Other factors such as the level of education among women, urban or rural dwelling, and occupation were associated with maternal knowledge of ODS (11). Strengthening ANC and multi-sectoral involvement by ensuring adequate education of the ODS may increase early recognition of complications and timely healthcare-seeking behaviors, and thereby significantly decreasing maternal mortality (12)-(13). Unfortunately, these and other factors are not known in many other countries of SSA which hinders the interventions towards preventing maternal mortality.

In Rwanda, 203 women die for every 100,000 live births (14). According to the most recent Rwanda Demographic Health Survey (2019-2020), the most common causes of maternal mortality are related to delay in seeking care (12–14). The importance of awareness and early recognition of ODS by women during pregnancy cannot be overstated. Evidence shows that women with better knowledge of ODS have improved healthcare seeking behavior than their counterparts with limited knowledge (3,9,15). Yet, the maternal knowledge of ODS in Rwanda is unknown limiting any interventions to decreasing maternal mortality. Accordingly, the purpose of the current study was to assess pregnant women’s knowledge of ODS during pregnancy, delivery, and postpartum period. The findings of this study are vital to policy makers to drive evidence-based care, hence leading to better maternal health outcomes.

### Objectives

To assess the maternal knowledge of ODS and associated factors during pregnancy delivery, and post-partum period among pregnant women attending ANC services in Kigali, Rwanda.

## METHODS

### Study design

This was a cross-sectional survey. To ensure the fullness of the report, we used the STROBE (Strengthening the Reporting of Observational Studies in Epidemiology) checklist (16).

### Study setting

The current study was conducted in the University Teaching Hospital of Kigali (CHUK), Muhima District Hospital, and 5 community health centers located in Nyarugenge District (Muhima, Bilyogo, Rugarama, Kabusunzu, and Rwampara).

### Participants, recruitment and sampling

Pregnant women of any gestational age, aged 18 years and older, receiving ANC at the study sites between September-December 2018 were eligible to participate in the study. We used convenient sampling of eligible pregnant women on the day of ANC appointments to the study sites. The consent was informed to participants and the essential points of the research were explained, including risks, benefits and the voluntary participation to help them making an informed choice. Those who agreed to participate signed the written informed consent and were enrolled in the study. 2 persons conducted interviews. The sample size was determined using a single population proportion formula (17), referring to the prevalence of knowledge of at least 3 ODS of 45.9%, with 95% confidence level, 5% margin of error reported by a study conducted in Ethiopia (17).

### Data source, variables, and measurements

We adapted the study questionnaire developed and initially tested in Ethiopia (18). The questionnaire consisted of 23 items asking information about maternal demographics, obstetrical history, time to the health facility, source of ODS information, and knowledge of ODS in the three categories (pregnancy, labor and birth, and immediate postpartum). To ensure face and content validity of the instrument in the local context, we removed the item regarding ethnicity, changed the religions, and income. Other items were also adjusted accordingly to the local context. A bilingual speaker of English and Kinyarwanda, and expert in translation helped translate the questionnaire to from English to Kinyarwanda. We tested the tool on 30 participants at Muhima health center and in Muhima District Hospital and there were no changes in the tool.

In exploring the factors associated with the outcome of interest (knowledge of ODS), the independent variables were gravidity, marital status, occupation, and income level.

### Data collection, management, and analysis

Data were collected from pregnant women attending ANC clinics between September and December 2018. The investigators (EU, UP, NU) and one trained research assistant (BJ) collected data by administering a 20-minute questionnaire. Each research participant was interviewed alone, and the data collector completed the questionnaire as the mother responded to the questions. The data was directly entered on the paper copy and later entered in SPSS 21 for cleaning and analysis.

A mother was classified to be knowledgeable if she had mentioned at least three ODS for each category (during pregnancy, during labor, and after delivery).

We used descriptive statistics (frequency, means, and standard deviations, and percentages) to report descriptive findings, and one-way Analysis of Variance (ANOVA) to assess the factors of maternal knowledge of ODS. The Fisher’s Exact Test was used to assess the association between knowledge of ODS and other variables for counts less than 5. The significance level was at 0.05.

## Ethical considerations

This study was approved by the Institutional Review Board (No317/CMHS/IRB/2018) of the College of Medicine and Health Sciences at the University of Rwanda and the CHUK Research and Ethics Committee (EC/CHUK/640/2018).

Participation in the study was voluntary. Pregnant mothers received the explanation of the study’s intent, what their participation will entail. Pregnant women who agreed to participate signed the consent form prior to initiating the interview. The collected data was kept confidential on a Password Protected laptop only accessible to other researchers within manuscript itself

## RESULTS

A total of 382 pregnant women met inclusion criteria and consented to participate in the study. The demographic characteristics are presented in (Table 1). The majority (67.8%) were aged 23-35 years, 44.5% completed secondary-level education, and 40.1% completed primary level education. The two frequent religions among participants were Protestantism (36.1%) and Catholicism (35.9%). 88.2% were married, and 58.0% were housewives with no other type of employment. 68.6% had less than 25, 000 Rwandan Francs (≈ $25USD) monthly income, and over two-thirds (69.4%) lived in the urban area. The participants’ obstetrical history indicated that 60.7% were pregnant for the first or second time, while 9.9% had had five or more pregnancies including the current pregnancy. Over half (59.9%) attended the first ANC visit in the first trimester and 57.6% completed at least three ANC visits in the previous pregnancy. The majority of participants (43.2%) needed more than 30 minutes to walk to the maternal health facilities; 33.5% walked 15 – 30 minutes, and 23.3% walked 15 minutes or less.

**Table 1.**
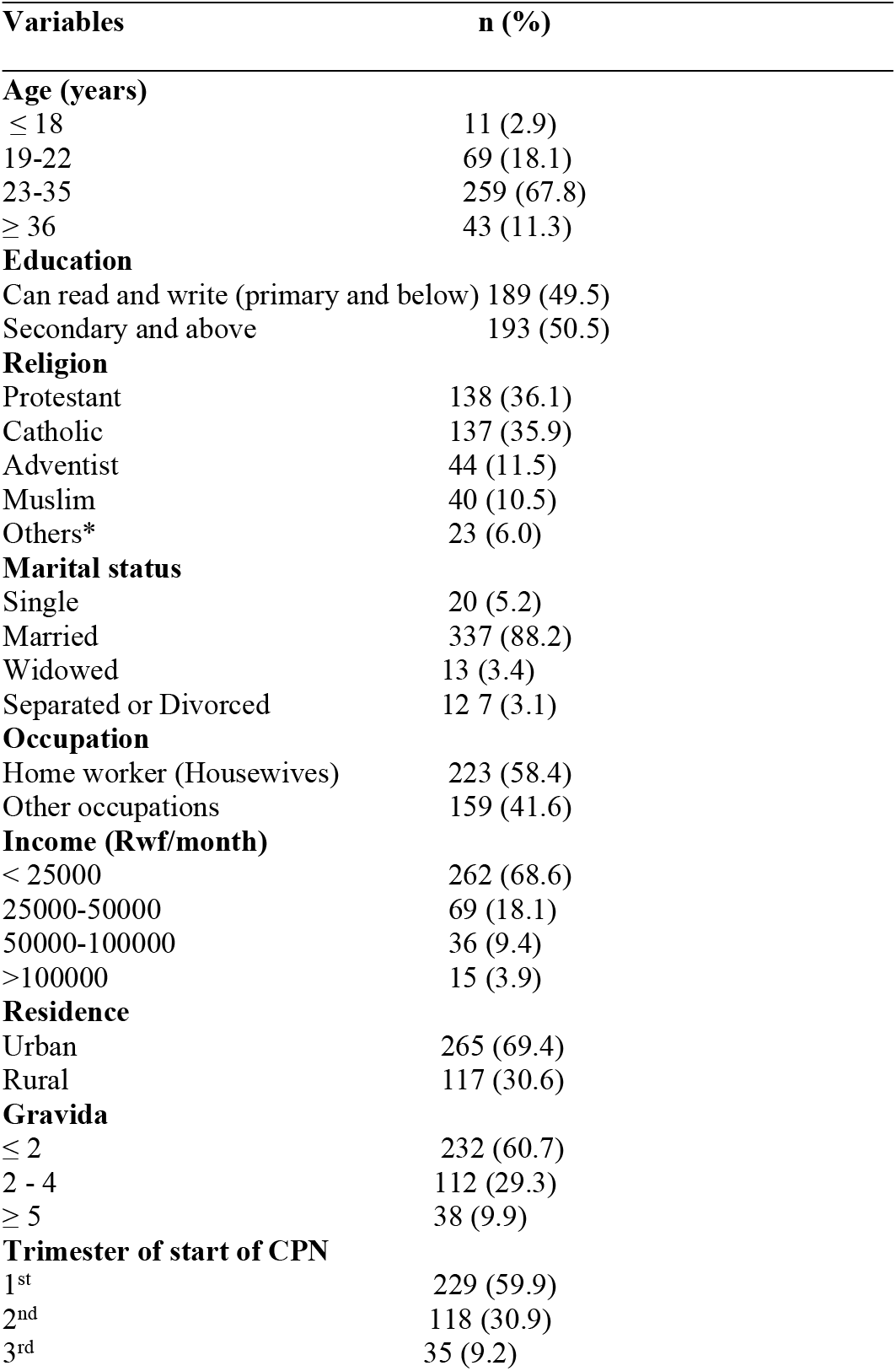
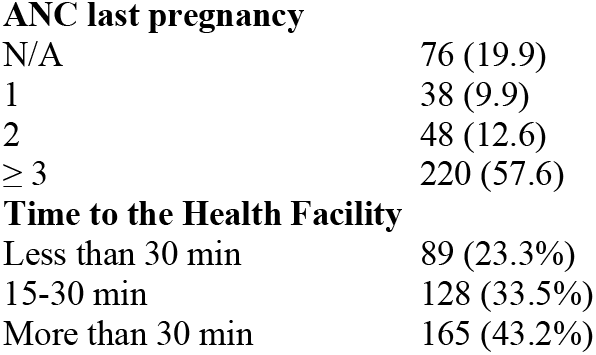
Demographic characteristics of participants (n=382)

| **Variables** | **n (%)** |
| --- | --- |
| **Age (years)** |  |
| ≤ 18 | 11 (2.9) |
| 19-22 | 69 (18.1) |
| 23-35 | 259 (67.8) |
| ≥ 36 | 43 (11.3) |
| **Education** |  |
| Can read and write (primary and below) | 189 (49.5) |
| Secondary and above | 193 (50.5) |
| **Religion** |  |
| Protestant | 138 (36.1) |
| Catholic | 137 (35.9) |
| Adventist | 44 (11.5) |
| Muslim | 40 (10.5) |
| Others* | 23 (6.0) |
| **Marital status** |  |
| Single | 20 (5.2) |
| Married | 337 (88.2) |
| Widowed | 13 (3.4) |
| Separated or Divorced | 12 (3.1) |
| **Occupation** |  |
| Home worker (Housewives) | 223 (58.4) |
| Other occupations | 159 (41.6) |
| **Income (Rwf/month)** |  |
| < 25000 | 262 (68.6) |
| 25000-50000 | 69 (18.1) |
| 50000-100000 | 36 (9.4) |
| >100000 | 15 (3.9) |
| **Residence** |  |
| Urban | 265 (69.4) |
| Rural | 117 (30.6) |
| **Gravida** |  |
| ≤ 2 | 232 (60.7) |
| 2 - 4 | 112 (29.3) |
| ≥ 5 | 38 (9.9) |
| **Trimester of start of CPN** |  |
| 1<sup>st</sup> | 229 (59.9) |
| 2<sup>nd</sup> | 118 (30.9) |
| 3<sup>rd</sup> | 35 (9.2) |

|  |  |  |
| --- | --- | --- |
| 514 | **ANC last pregnancy** |  |
| 515 | N/A | 76 (19.9) |
| 516 | 1 | 38 (9.9) |
| 517 | 2 | 48 (12.6) |
| 518 | ≥ 3 | 220 (57.6) |
| 519 | **Time to the Health Facility** |  |
| 520 | Less than 30 min | 89 (23.3%) |
| 521 | 15-30 min | 128 (33.5%) |
| 522 | More than 30 min | 165 (43.2%) |

Overall, the women’s knowledge of at least three ODS during pregnancy, labor and delivery, and post-partum was 56.6%, 9.2% and 17.5%, respectively (Figure 1). Participant’s knowledge of ODS during pregnancy, labor and birth, and immediate postpartum are presented in Table 2. 70.9% were aware that vaginal bleeding is a ODS during pregnancy, and about half (51.3%) were aware of severe abdominal pain as an ODS. Participants were less knowledgeable of the ODS of labor and birth, with only a third (35.6%) being aware of vaginal bleeding. A limited proportion (11.8% and 9.2%) were aware of labor lasting greater than 12 hours, and a delay of 30 minutes with placenta delivery as ODS, respectively. Half (52.4%) were aware of vaginal bleeding post-partum to be an ODS, and a limited proportion (13.6%) recognized that severe weakness and malodorous vaginal discharge post-partum were ODS.

**Table 2.**
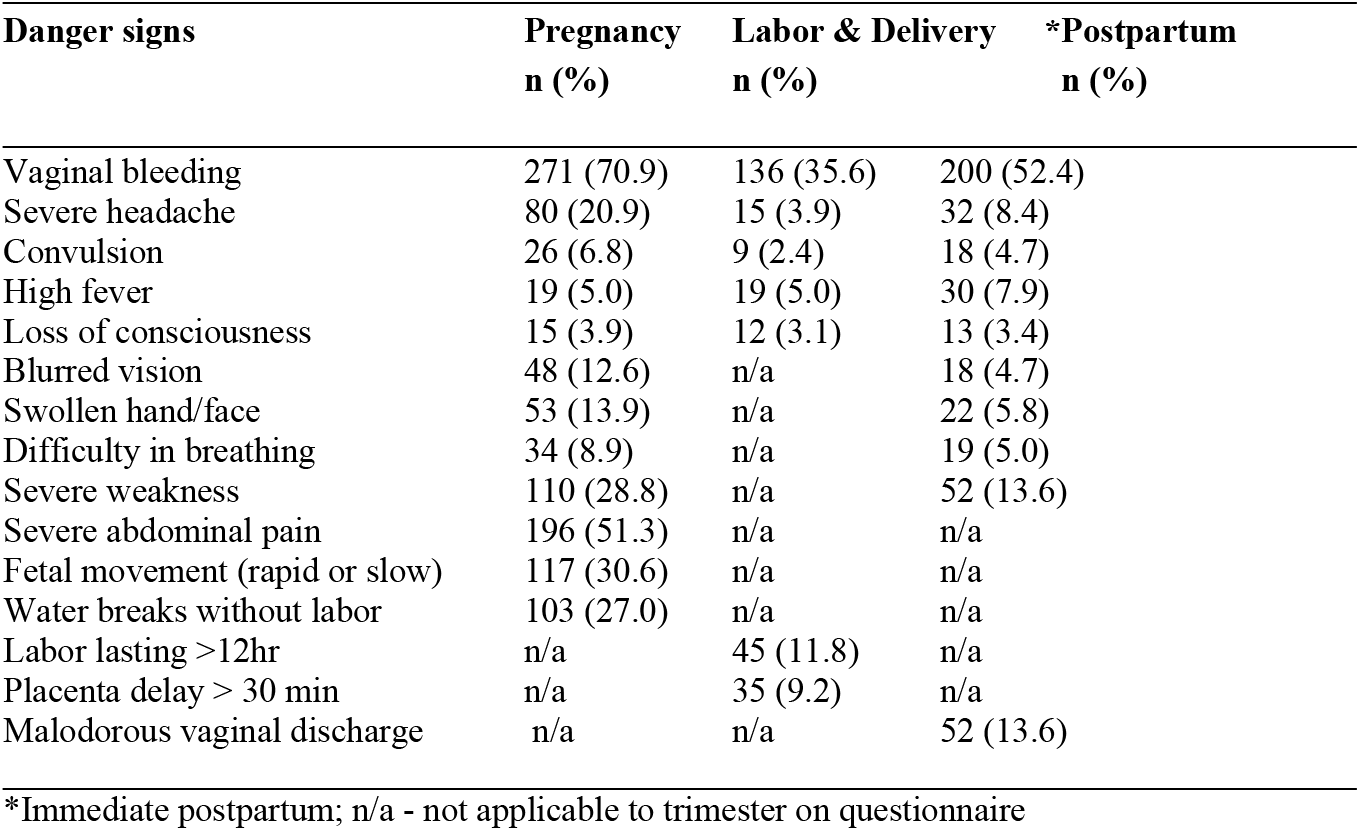
Knowledge of ODS during pregnancy, labor and birth, and immediate postpartum (n = 382)

**Figure 1.**
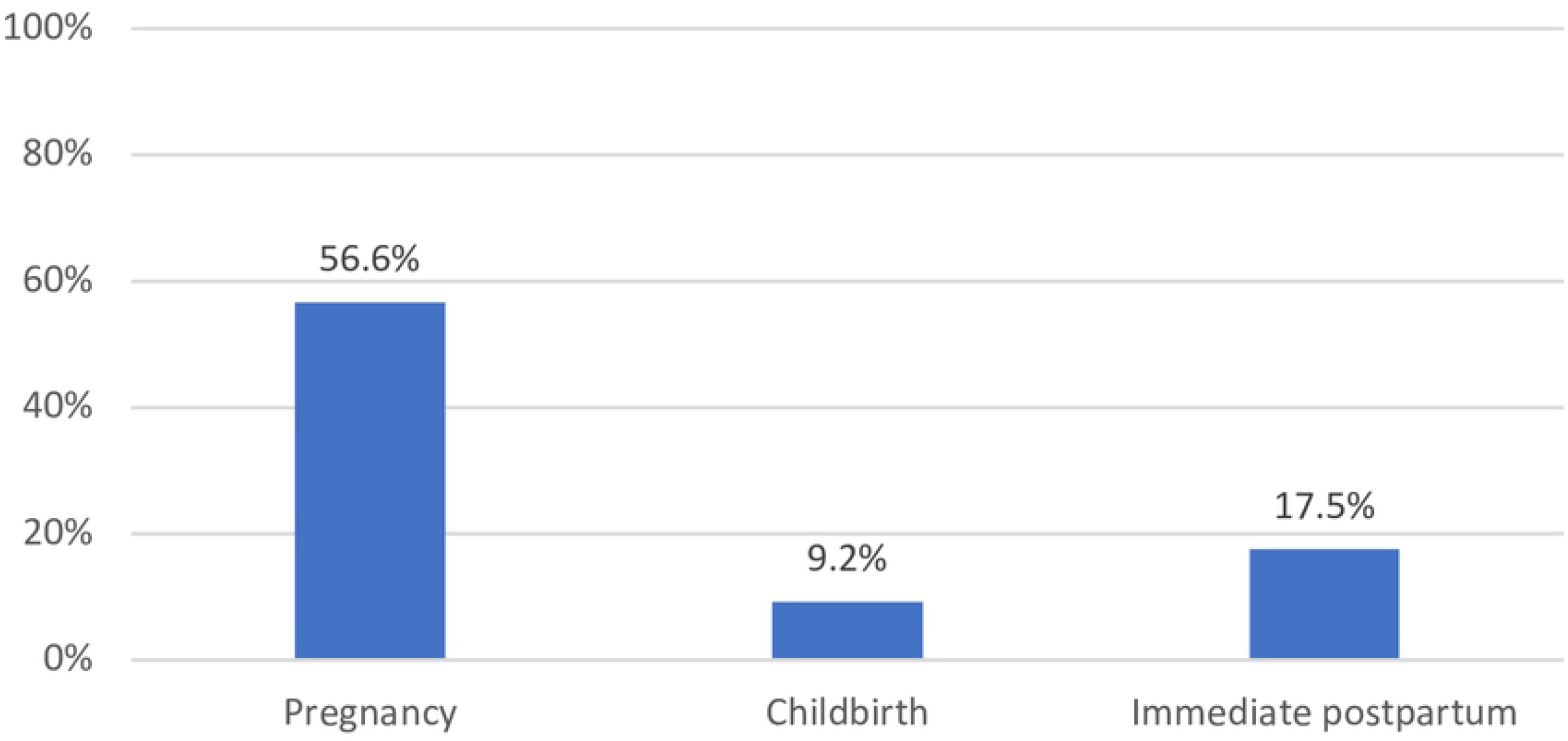
Overall Knowledge of at least 3 obstetric danger signs.

327 women responded to the question about the source of ODS information. 46.1% of them got information from nurses and midwives, 31.2% from fellow mothers, and 20.7% from community health workers (Figure 2).

**Figure 2.**
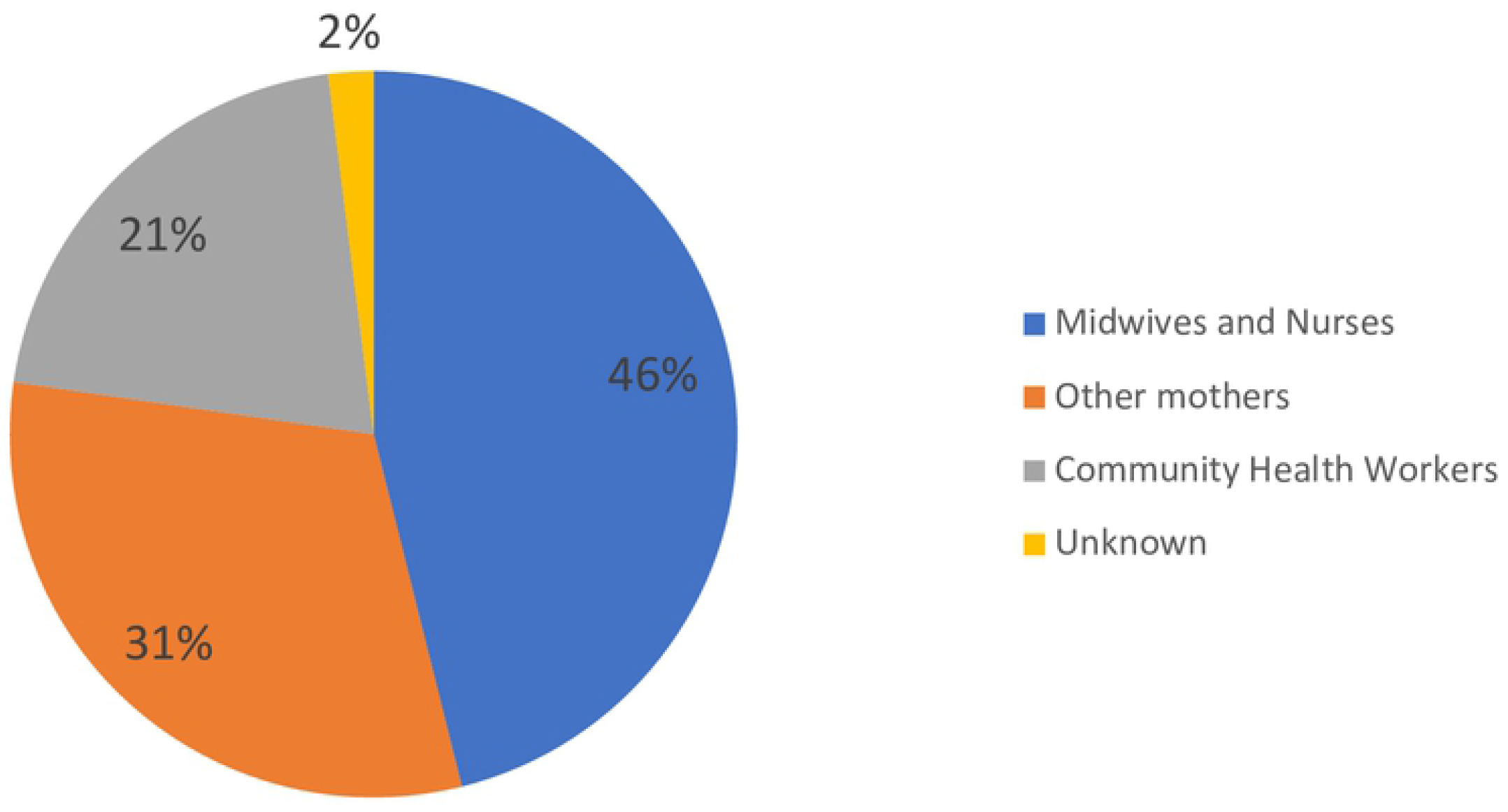
Source of information about obstetric danger signs (n=327)

The knowledge of ODS during pregnancy was associated with gravidity (p = 0.015), marital status (p = 0.034), occupation (p = 0.003), and income (p = 0.044). During labor and birth, there was a statistical significance between ODS knowledge and marital status (p = 0.04). During the immediate postpartum period, there was a statistical significance between ODS knowledge and occupation (p = 0.05). All other variables had no statistical significance. (Table 3).

**Table 3.**
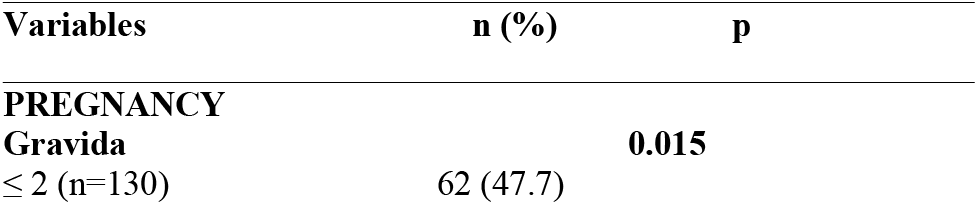
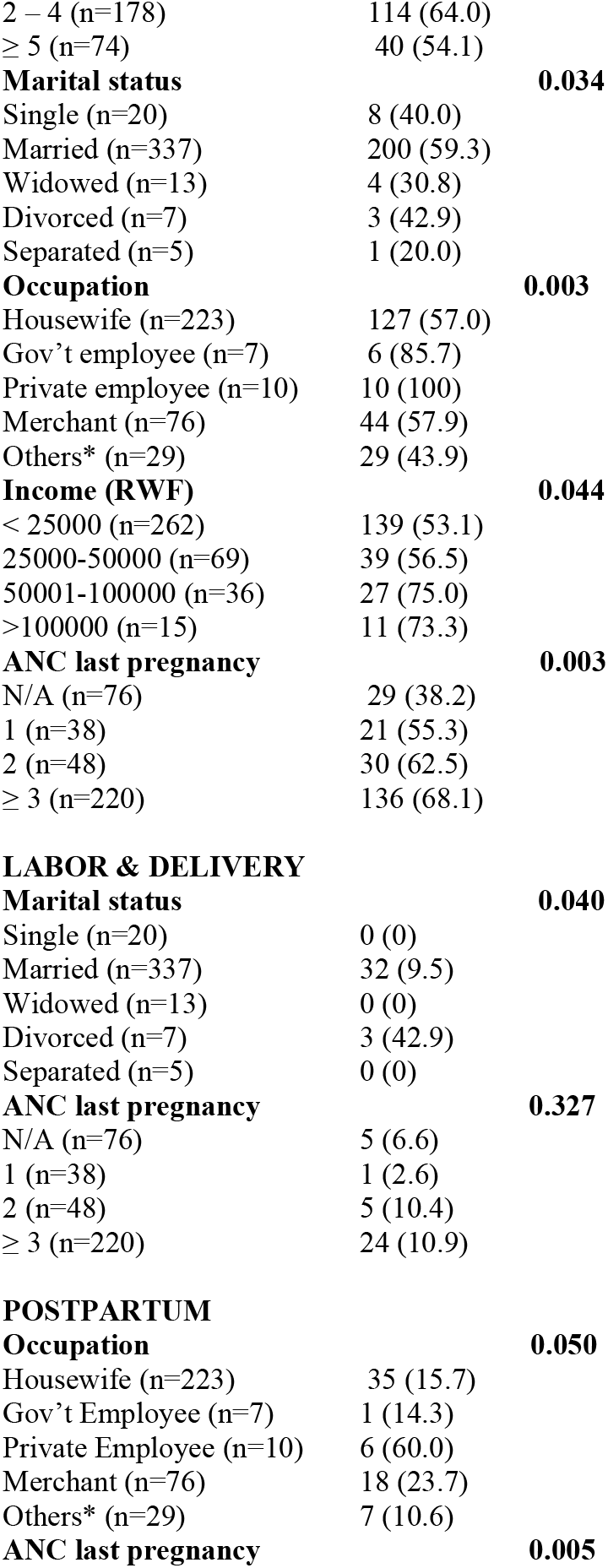
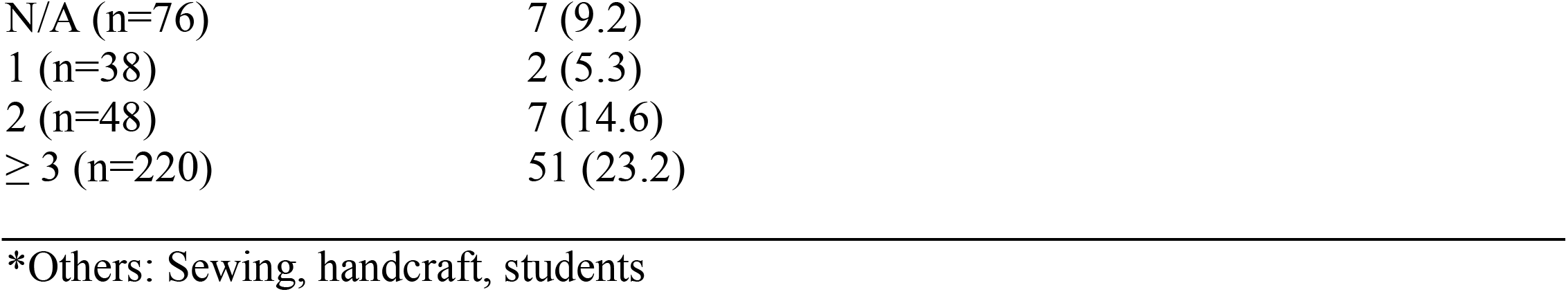
Factors associated with the knowledge of ODS during pregnancy, labor and delivery, and postpartum.

| **Variables** | **n (%)** | **p** |
| --- | --- | --- |
| **PREGNANCY** |  |  |
| **Gravida** |  | **0.015** |
| ≤ 2 (n=130) | 62 (47.7) |  |

|  |  |  |  |
| --- | --- | --- | --- |
| 560 | 2 – 4 (n=178) | 114 (64.0) |  |
| 561 | ≥ 5 (n=74) | 40 (54.1) |  |
| 562 | **Marital status** |  | **0.034** |
| 563 | Single (n=20) | 8 (40.0) |  |
| 564 | Married (n=337) | 200 (59.3) |  |
| 565 | Widowed (n=13) | 4 (30.8) |  |
| 566 | Divorced (n=7) | 3 (42.9) |  |
| 567 | Separated (n=5) | 1 (20.0) |  |
| 568 | **Occupation** |  | **0.003** |
| 569 | Housewife (n=223) | 127 (57.0) |  |
| 570 | Gov't employee (n=7) | 6 (85.7) |  |
| 571 | Private employee (n=10) | 10 (100) |  |
| 572 | Merchant (n=76) | 44 (57.9) |  |
| 573 | Others* (n=29) | 29 (43.9) |  |
| 574 | **Income (RWF)** |  | **0.044** |
| 575 | < 25000 (n=262) | 139 (53.1) |  |
| 576 | 25000-50000 (n=69) | 39 (56.5) |  |
| 577 | 50001-100000 (n=36) | 27 (75.0) |  |
| 578 | >100000 (n=15) | 11 (73.3) |  |
| 579 | **ANC last pregnancy** |  | **0.003** |
| 580 | N/A (n=76) | 29 (38.2) |  |
| 581 | 1 (n=38) | 21 (55.3) |  |
| 582 | 2 (n=48) | 30 (62.5) |  |
| 583 | ≥ 3 (n=220) | 136 (68.1) |  |
| 584 |  |  |  |
| 585 | **LABOR & DELIVERY** |  |  |
| 586 | **Marital status** |  | **0.040** |
| 587 | Single (n=20) | 0 (0) |  |
| 588 | Married (n=337) | 32 (9.5) |  |
| 589 | Widowed (n=13) | 0 (0) |  |
| 590 | Divorced (n=7) | 3 (42.9) |  |
| 591 | Separated (n=5) | 0 (0) |  |
| 592 | **ANC last pregnancy** |  | **0.327** |
| 593 | N/A (n=76) | 5 (6.6) |  |
| 594 | 1 (n=38) | 1 (2.6) |  |
| 595 | 2 (n=48) | 5 (10.4) |  |
| 596 | ≥ 3 (n=220) | 24 (10.9) |  |
| 597 |  |  |  |
| 598 | **POSTPARTUM** |  |  |
| 599 | **Occupation** |  | **0.050** |
| 600 | Housewife (n=223) | 35 (15.7) |  |
| 601 | Gov't Employee (n=7) | 1 (14.3) |  |
| 602 | Private Employee (n=10) | 6 (60.0) |  |
| 603 | Merchant (n=76) | 18 (23.7) |  |
| 604 | Others* (n=29) | 7 (10.6) |  |
| 605 | **ANC last pregnancy** |  | **0.005** |
| 606 | N/A (n=76) | 7 (9.2) |
| 607 | 1 (n=38) | 2 (5.3) |
| 608 | 2 (n=48) | 7 (14.6) |
| 609 | $\geq 3$ (n=220) | 51 (23.2) |

611 \*Others: Sewing, handcraft, students

## DISCUSSION

In the current study, we explored the maternal knowledge of ODS during pregnancy, labor and delivery, and immediate post-partum period. Our results indicate that 56.6% had knowledge of at least three ODS during pregnancy. The findings about maternal knowledge of ODS vary across studies and countries but a similar pattern in SSA is that women are less knowledgeable. Our findings fall in the same pattern. Studies conducted in Ethiopia, Uganda, and South Africa reported that maternal knowledge of at least 3 ODS was 71.3%, 31.3% and 67.6% respectively (6,8,19). It is however important to highlight that the reporting of findings about knowledge of ODS is rather non-uniform. Some studies define maternal knowledge as knowing 3 ODS, others 4 or even 5. This renders the comparability of findings ambiguous necessitating scrutiny. In anyway, the maternal knowledge of ODS is the first step in identifying potential complications and prompts women early care seeking, hence appropriate management, and timely referral if necessary. Timely identification and good-quality management of obstetrical complications can contribute to reducing the burden of maternal mortality and associated long-term complications to both the mother and newborn (20).

The women’s knowledge of ODS during pregnancy is vital. Worldwide, the leading causes of maternal mortality in pregnancy are hemorrhage, maternal sepsis, and hypertensive disorders (21). The signs and symptoms of these three include; vaginal bleeding for haemorrhage; fever for sepsis; and swollen limbs, face, severe headache, and blurred vision for hypertensive disorders [15]. On the positive side, across all categories (pregnancy, labor and delivery, and immediate postpartum), vaginal bleeding was the most frequently reported ODS. Similarly, several studies in Uganda, Tanzania and Ethiopia reported that high women‘s awareness of vaginal bleeding as an ODS, is due to blood-red being symbolized as a danger in African cultures; thus, it is easily recognized by women (6,7,26–29,8,9,11,18,22–25). These findings are encouraging given that 33% of all maternal deaths are caused by post-partum haemorrhage [18]. On the other hand, fever is an important sign of sepsis yet was one of the least mentioned ODS despite the risk of complications to both the mother and the baby. In Kenya, fever was the third common ODS mentioned by women (30). Despite the hypertensive disorders being one of the important causes of maternal deaths, results from studies reported low awareness of headache as an ODS. In the current study only 20% of women knew that severe headache during pregnancy is an ODS, whereas in Tanzania the awareness was 44%(9). The study in Tanzania included women who had delivered within two years, and with majority having more than 2 previous pregnancies which could explain a relatively higher awareness compared to the current study. The factors associated with the maternal knowledge of ODS are relatively consistent across studies in SSA. Similar to our findings, a study conducted in Tanzania reported maternal knowledge of ODS to be associated with educational status, occupation, attendance to the ANC services, and received counselling on ODS (9). In Kenya, the statistically significant factor of knowledge of ODS was the attendance of antenatal care services (30). The pregnancy experience related factors of knowledge of ODS such as a woman’s gravidity, maternal age, can be expected. The experience of becoming pregnant and utilizing health services over and over make women with higher number of past pregnancies – usually those with relatively higher age become more knowledgeable than their inexperienced and younger counterparts. Such findings point fingers to the potential value of structured peer education among pregnant women at the ANC sessions or in the community. Peer education can be influential in mothers changing or adopting certain health behaviors(31). Peer education method was effective in Tanzania helping young mothers become knowledgeable of ODS(32). The young mothers who received the intervention of learning ODS showed higher care seeking behaviors than the comparison group.

The understanding of other factors of maternal knowledge of ODS is necessary especially for the development of interventions for improving maternal and neonatal outcomes. The current and several other studies reported that married women are likely to be more knowledgeable on ODS than their unmarried counterparts. Married women were more likely to be knowledgeable than single or divorced women, similar to results from a study in Kenya (30). In the Rwanda culture, pregnancy among married women is a source of pride which could provide women the ease and motivation of learning more about ODS. Better knowledge among married women could also be associated with husbands being more involved in the pregnancy experience. Women with supportive husbands are more likely to feel better about their pregnancy and want to be protective of themselves and their babies(33). A study conducted Nigeria exploring men’s knowledge of ODS found it to be very low, especially dangers signs during postpartum period (34). As the authors recommended, higher husbands’ knowledge of ODS could improve maternal and newborn health outcomes because husbands are usually the closest to mothers to help detect ODS. While multiple studies advocate for and highlight the importance of men’s involvement in learning and early detection of ODS (34)(35), a qualitative study in Ghana reported that men’s increased involvement would weaken an aspect of a women’s life where she has traditionally had some authority (35). Amidst this controversy, it is important to remember that childbearing should be a shared decision between the couple which makes both parties’ involvement necessary.

Employment status of mothers is another factor of maternal knowledge of ODS. There more than one way in which employment helps mothers be better knowledgeable. First, employed women are likely to be more educated which in one way could explain better knowledge of ODS. Second, employment is a source of income which could help easier utilization of quality healthcare services [9, 20], hence an avenue to be educated on ODS. In a study conducted in Ethiopia, employed women faced fewer barriers than unemployed women and women who worked as self-employed were about five times more knowledgeable of ODS than houseworkers (36). Employed and educated people are likely to be socially connected to people who can provide health information which could help alert women on ODS better than their unemployed counterparts. Women’s employment also increases the family income, allowing more opportunities to obtain health information such as access to other resources such as the radio, internet, and television (36).

### Limitations

This study used a cross-sectional design hence we cannot learn the trend in maternal knowledge of ODS. Second the setting of the study was Kigali – a capital city of Rwanda, therefore the results may not be generalizable to other regions especially the rural population. It was an observational study, and therefore we cannot conclude a causal relationship between ODS knowledge and explanatory variables. The study was conducted when the women were pregnant, so familiarity with ODS in the other perinatal categories (birth and postpartum) may have been less important at the time of data collection. The cultural and religious factors may also influence knowledge of ODS though we did not collect this information.

### Recommendations

Based on our findings, cooperation among different ministries, organizations, and private sectors is needed to increase educational promotion during the perinatal period. A clear policy on ANC with a structured plan that includes patient education at each visit would likely increase knowledge acquisition and retention. Sensibilization of ANC content via the media, meetings and campaigns would likely decrease the number of those who start ANC late and miss important ODS education. Also, education on the danger signs for newborns, must include videos during ANC and vaccination clinics. Encouraging the involvement of husbands in all perinatal visits at health centers and hospitals has the potential to improve outcomes, as well as encouraging legal marriage.

### Conclusion

This study assessed pregnant women’s knowledge of obstetrical danger signs during pregnancy, labor and delivery, and immediate postpartum. Maternal knowledge was found to be very low especially the knowledge of ODS during labor and delivery, and immediate post-partum period even though most maternal deaths occur during these two periods. marriage, and occupation were significantly associated with maternal knowledge of obstetrical danger signs. It is essential to emphasize on the quality of ANC contacts ensuring that mothers (and where possible their spouses) are equipped with the knowledge they require to quickly notice danger signs and seek care. Additional avenues for educating women need to be identified such as in the community by the help of Community Health Workers, during prenuptial counseling, postpartum encounters and so on. Effective strategies such as peer education, videos could be further experimented. Health care providers need to be well educated on the importance of sharing information with pregnant women and provided educational tools. This study could serve as a baseline for further research to explore knowledge of ODS not only among pregnant women but also among healthcare providers in different parts of Rwanda.

## Data Availability

In the online submission form, you indicated that "The collected data was kept confidential, on a Password Protected laptop Only accessible to the research team prior to the deidentification but provided the dataset in .csv format and uploaded it as a supplementary file" All PLOS journals now require all data underlying the findings described in their manuscript to be freely available to other researchers, either 1. In a public repository, 2. Within the manuscript itself, or 3. Uploaded as supplementary information.

## Abbreviation

ANC: Antenatal Care
ODS: Obstetrics Danger Signs
DH: District Hospital
HC: Health Center
HEW: Health Expert Worker
MMR: Maternal Mortality Rate
MDG: Millennium Development Goals
PI: Principal Investigator
RDHS: Rwanda Demographic and Health Survey
SPSS: Statistical Package for the Social Sciences
UTHK: University Teaching Hospital of Kigali
WHO: World Health Organization

## Declaration

- **Availability of data and materials:** Data sheet available here
- **Competing interests**: The authors have declared that no competing interests exist
- **Funding**: The authors received no specific funding for this work
- **Authors’ contributions:**

Conception: Emmanuel Uwiringiyimana

Data collection and Analysis: Emmanuel Uwiringiyimana Albert Nsanzimana, Neophyte Uhawenayo, Pacifique Ufitinema, Janviere bayizere

Methodology and design: Emmanuel Uwiringiyimana, Emery Manirambona, Samuel Byiringiro

**Writing – Original Draft Preparation:** Emmanuel Uwiringiyimana, Emery Manirambona, Samuel Byiringiro, Albert Nsanzimana, Neophyte Uhawenayo, Pacifique Ufitinema, Janviere bayizere

**Writing – Review & Editing for intellectual content:** Patricia J. Moreland, Pamela Meharry, Diomede Ntasumbumuyange

**Validation and supervision:** Patricia J. Moreland, Pamela Meharry, Diomede Ntasumbumuyange

## Acknowledgements

We would like to thank the midwives and health administrators at the health facilities for their cooperation with the data collection, Daniel Bogale, Assistant professor who gave us the structured questionnaire, and Becky White who helped in structuring the title of this research project.

